# Evaluation of recombinant nucleocapsid and spike proteins for serological diagnosis of novel coronavirus disease 2019 (COVID-19)

**DOI:** 10.1101/2020.03.17.20036954

**Authors:** Pingping Zhang, Qi Gao, Tang Wang, Yuehua Ke, Fei Mo, Ruizhong Jia, Wanbing Liu, Lei Liu, Shangen Zheng, Yuzhen Liu, Luping Li, Yao Wang, Lei Xu, Kun Hao, Ruifu Yang, Shiyue Li, Changqing Lin, Yong Zhao

**Affiliations:** State Key Laboratory of Pathogen and Biosecurity, Beijing Institute of Microbiology and Epidemiology, Beijing 100071, P. R. China; Beijing Key Laboratory of POCT for Bioemergency and Clinic (No. BZ0329), Beijing 100071, P. R. China; Beijing Hotgen Biotechnology Inc., Beijing 100071, P. R. China; Wuhan University School of Health Sciences, P. R. China; Huoshenshan Hospital, Wuhan, P. R. China; Center for Clinical Laboratories, Affiliated Hospital of Guizhou Medical University, Guiyang 550004, P. R. China; Department of Basic Clinical Laboratory Medicine, School of Clinical Laboratory Science, Guizhou Medical University, Guiyang 550004, P. R. China; General Hospital of Central Threater Command of the PLA, Wuhan, P. R. China; Shijiazhuang Fifth Hospital, Shijiazhuang 050023, P. R. China; The Sixth People’s Hospital of Shenyang, P. R. China; Department of Clinical Laboratory, Peking Union Medical College Hospital, Peking Union Medical College, Chinese Academy of Medical Sciences, P. R. China

**Keywords:** HCoV-19 (SARS-CoV-2), Recombinant protein, Eukaryotic expression, Serological diagnosis, Colloidal gold immunochromatography assay (GICA)

## Abstract

**Background:** The colloidal gold immunochromatography assay (GICA) is a rapid diagnostic tool for novel coronavirus disease 2019 (COVID-19) infections. However, with significant numbers of false negatives, improvements to GICA are needed.

**Methods:** Six recombinant HCoV-19 nucleocapsid and spike proteins were prepared and evaluated. The optimal proteins were employed to develop a sandwich-format GICA strip to detect total antibodies (IgM and IgG) against HCoV-19. GICA’s performance was assessed with comparison of viral RNA detection.

**Results:** Recombinant HCoV-19 proteins were obtained, including three prokaryotically expressed rN, rN1, rN2 nucleocapsid proteins, and three eukaryotically expressed rS1, rS-RBD, rS-RBD-mFc spike proteins. The recombinant proteins with the highest ELISA titers (rS1 and rS-RBD-mFc) against coronavirus-specific IgM and IgG were chosen for GICA development. The GICA has a sensitivity and specificity of 86.89% (106/122) and 99.39% (656/660), respectively. Furthermore, 65.63% (21/32) of the clinically confirmed but RT-PCR negative samples were GICA positive.

**Conclusions:** The eukaryotically-expressed spike proteins (rS1and rS-RBD-mFc) are more suitable than the prokaryotically expressed nucleocapsid proteins for HCoV-19 serological diagnosis. The GICA sandwich used to detect total antibodies is a powerful complement to the current standard RNA-based tests.

## Introduction

The recent coronavirus disease 2019 (COVID-19) outbreak has been classified as a global pandemic on March 12, 2020, which continues to pose a great threat to global public health. The disease is caused by a novel coronavirus, called 2019-nCoV by the World Health Organization at the beginning of the outbreak, and latterly, severe acute respiratory syndrome coronavirus 2 (SARS-CoV-2) by the ICTV (International Committee on Taxonomy of Viruses) [1]. However, this name is prone to confusion with SARS-CoV, which emerged in 2003, and a suggestion for a distinct name was proposed, human coronavirus 2019 (HCoV-19) [2]. This novel virus is potentially more transmissible than SARS-CoV and other coronavirus [3], making early diagnosis of it important for clinical treatment and disease control. However, the current nucleic acid testing approach for HCoV-19 [4] carries a negligible false-negative risk [5]. Serological assays are supposedly a powerful approach for achieving timely diagnosis of COVID-19, as is nucleic acid testing, especially for patients with undetectable viral RNA [6]. The serological assays used for early diagnosis are mainly based on detecting specific antibodies against HCoV-19 in a patient’s serum and include the enzyme-linked immunosorbent assay (ELISA), the colloidal gold immunochromatography assay (GICA) and the chemiluminescence assay.

To develop a reliable serological assay, it is important to obtain suitable HCoV-19 antigens or related recombinant proteins. HCoV-19 is a β family coronavirus, with a spike protein (S protein), envelope protein (E), membrane protein (M), and nucleocapsid protein (N protein). Among them, the N-protein is the most abundant, relatively conservative protein in coronaviruses; thus, it is often used as a diagnostic antigen [7]. Our previous study showed that the antigenicity of the COOH terminus is higher than that of the NH_2_ terminus in the SARS-CoV N protein, and that the former N protein fragment may have the same antigenicity as that seen with the full-length N protein [8]. In contrast, the S protein is the common target when designing vaccines based on neutralizing antibodies. It contains a receptor binding domain (S-RBD) in the S1 subunit, which mediates receptor binding and membrane fusion [9, 10]. Recently, S-RBD from HCoV-19 was reported to have higher affinity than SARS-CoV to angiotensin-converting enzyme 2, and also to lack cross-reactivity with monoclonal antibodies against S-RBD from SARS-CoV [3], highlighting the potential value of the S protein for diagnosing HCoV-19. As a transmembrane protein with hydrophobic regions, the S protein is better prepared using a eukaryotic expression system.

In the present study, we prepared six recombinant proteins based on the reported HCoV-19 sequence (GenBank accession MN908947) [11]. Three recombinant N proteins were obtained by prokaryotic expression, including the full-length recombinant N protein (rN), NH_2_ terminal (rN1) protein fragments, and COOH terminal (rN2) fragments from this protein. Three recombinant S proteins were also obtained by eukaryotic expression, including an S1 domain (rS1) fragment, the receptor binding domain (rS-RBD), and S-RBD ligated to the Fc fragment from mouse (rS-RBD-mFc). The recombinant proteins were evaluated using indirect ELISA. Two recombinant proteins with the highest ELISA titers (rS1and rS-RBD-mFc) were chosen to develop a sandwich-format GICA strip to detect total antibodies (IgM and IgG) against HCoV-19. The performance of the GICA strip, which was evaluated with 814 clinical samples, showed a high detection sensitivity (86.89%) and specificity (99.39%) with samples from COVID-19-infected patients.

## Materials and Methods

### Ethics statement

This study was approved by the Medical Ethical Committee of Peking Union Medical College Hospital (approval number 002285), General Hosptical of Central Threater Command of the PLA (approval number 2020-003-1), and Shijiazhuang Fifth Hospital (approval number 2020-002). Written informed consent from all the patients was obtained, and all records and information on the patients were anonymized.

### Materials

DNA polymerase (2×Pfu MasterMix) was purchased from Beijing TransGen Biotech (Beijing, China). T4 DNA ligase and Gibison Assembly kit were from New England BioLabs Inc., (Ipswich, England). Eukaryotic vectors H293 and H293-Fc, which were used for transient expression, were obtained from the Laboratory of Protein Project, Beijing Institute of Biotechnology, China. FreeStyle™ 293 expression medium, Opti-MEM^®^ I (1×), a reduced serum medium, and 293 fectin™ reagents were purchased from Invitrogen Inc., (CA, Carlsbad, USA). The Unique CDSystem for protein purification was from Suzhou Inscinstech Co., Ltd. (Suzhou, China). The DNA extraction/purification kit was from Beijing TransGen Biotech. Primer construction and sequencing work were conducted by Beijing Tianyi Huiyuan Biotech Ltd., and Beijing Biomed Biotech Ltd., respectively. GICA nitrocellulose (NC) membranes were obtained from MilliporeSigma. (Saint Louis, MO, USA), glass fiber and absorbent pads were from Shanghai Kinbio Tech. Co., Ltd. (Shanghai, China).

The gene sequences of the recombinant proteins were all derived from the RNA sequence of HCoV-19 strain from Wuhan (GenBank accession MN908947). The nucleotide positions of the gene sequences are rS1 (21602-23584), rS-RBD (22514-23311), rN (28274-29530), rN1 (28274-28900), and rN2 (28901-29530). The N, N1, and N2 gene cloning vectors were constructed by General Biosystems Co., Ltd (Anhui, China). The S1 DNA sequence was optimized and synthesized by GenScript Co., Ltd (Nanjing, China). The vector and bacterial strains for prokaryotic expression were pET28a, *E. coli* Rosetta or BL21(DE3).

### Gene subcloning, eukaryotic expression vector construction, and protein expression and purification of rS1, rS-RBD-mFc and rS-RBD

The full coding region, which was obtained by overlapping extension PCR using primers containing restriction enzyme recognition sites (Table 1), was ligated to HEK293 vectors after digestion. The recombinant plasmids were transformed into *E. coli* DH5α, and bacterial colonies were selected on Luria-Bertani (LB) agar containing ampicillin. Positive colonies were cultured in 500 μL LB liquid medium for 2–4 h, and the resultant plasmids were extracted, PCR-verified, and sequenced. Positive recombinant plasmids (210 μg each) and liposomes (280 μL) were diluted in 7 mL of opti-MEM medium for 5 min, respectively, mixed for 30 min and H293 cells (1.2× 10^6^ /mL) were added. After culturing (120 rpm, 37°C with 5% CO_2_, 3–4 days), the cell supernatants collected by centrifugation were purified with a 0.44-μm filter and the Unique CDSystem chromatography workstation. After column Protein A balancing (10 column volumes) with phosphate-buffered saline (PBS), the cell supernatants were placed under a flow rate of 2.0 ml/min, and then washed and eluted with PBS (five column volumes) and citric acid buffer (pH 3.0) to collect the purified protein. For desalination, a 1/3 sample volume was applied to a G25 column pre-balanced with PBS (5 column volumes), and each protein was collected after column washes with PBS. Their purities were confirmed by sodium dodecyl sulfate–polyacrylamide gel electrophoresis (SDS-PAGE).

### Gene subcloning, expression vector construction, protein expression and purification of rN, rN1 and rN2

The N protein’s full coding region was PCR amplified using primers containing restriction enzyme recognition sites (Table 1). The restricted amplicons were ligated into pET28a or pET32a using T4 DNA ligase. The recombinant plasmids were transformed into *E. Coli* Rosetta or BL21 (DE3), and the bacterial colonies were selected on LB agar containing kanamycin. Recombinant plasmids in the bacterial colonies were extracted and confirmed as authentic by DNA sequencing. Protein expression was induced for 5 h in positive bacterial colonies with 0.5 mM isopropyl-D-thiogalactopyranoside (IPTG) at a starting optical density (OD) of 0.6. After centrifugation, the supernatants and precipitation products from the lyzed bacteria were analyzed by SDS-PAGE. The rN, rN1 and rN2 recombinant proteins were produced by the bacteria harboring the recombinant plasmids via IPTG induction in 2 L of LB medium (20°C, 200 x g, 10 h), and the bacteria were harvested and lyzed by ultrasonication (300 W, 30 min). The supernatants were clarified by centrifugation (10,000 × g, 20 min), filtrated (0.4-μm filter membrane), and run through a 3 mL Ni Sepharose column with 30 ml lysis buffer, and then washed extensively with PBS containing imidazole (20–80 mM gradient) to remove non-specific proteins. The target proteins were eluted with PBS containing 250 mM imidazole.

**Table 1.**
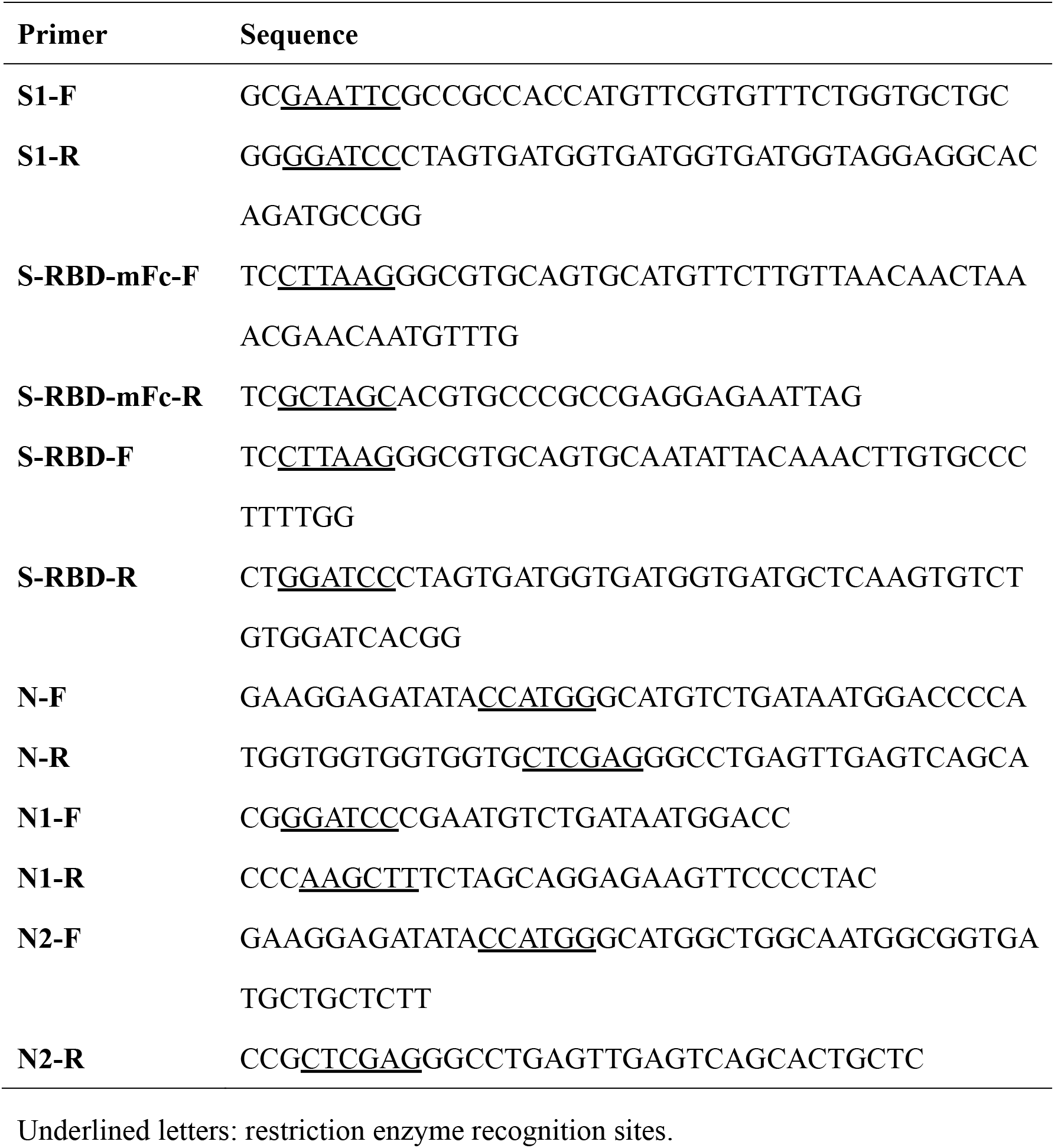
Subcloning primers for preparation of the recombinant proteins.

### Preliminary evaluation of recombinant proteins by ELISA

The recombinant proteins were initially evaluated by indirect ELISA. After coating the wells with the prepared recombinant proteins, 50μL of serially diluted human samples were added to the wells and mixed with 50μL of horseradish peroxidase (HRP)-labeled goat anti-human IgM or IgG, followed by incubation (37°C, 60 min). After the solution was removed and the wells had been washed with PBS buffer containing Tween-20 five times, 50 μL of tetramethylbenzidine substrate was added to the wells in the plate for 15 min. OD values were measured on a microplate reader (450/630 nm), and recombinant proteins with high OD values for COVID-19 patient serum and low OD reads for the negative control serum were used for all further experiments.

ELISA plates coated with the six recombinant proteins were first tested with seven negative sera from healthy people. Their average values plus two-fold standard deviations (MEAN+2SD) were used as the cutoffs. However, they were not completely equivalent for each protein and some were set at 0.2 when the calculated values were below 0.2. Sera from two patients with COVID-19 were diluted at ratios of 1:800, 1:400 and 1:100 for IgM detection, and ratio of 1:80, 1:40 and 1:20 for IgG detection. The average value (MEAN) from the negative serum samples was set as the background value, and the lowest antibody titer with a value higher than the cutoff was set as the sensitivity level.

### Preparing the GICA strip with recombinant proteins for antibody detection

Colloidal gold suspension was prepared by reducing gold chloride with citrate. The colloidal gold was conjugated to recombinant protein rS-RBD. Briefly, 1 mg of rS-RBD was added to 100 mL of the colloidal gold suspension. After a 30 min reaction, conjugation was blocked using 10 mL of 10% bovine serum albumin (BSA) for 15 min. The colloidal gold conjugate collected (centrifugation at 12,000 rpm, 30 min, 4°C) was resuspended in PBS containing 0.1% BSA and 0.1% Tween-20.

The colloidal gold and rS-RBD (0.5 mg/mL) conjugate was applied to a conjugate pad (glass fiber) (30 μL/cm, dried at 37 °C, 3 h). Using a dispenser (XYZ3000; BioDot, Irvine, CA), rS1 and the secondary polyclonal antibody (2 mg/mL) were coated onto the nitrocellulose membrane as the test and control lines, respectively, at a dispensing rate of 1.0 μL/cm. The membrane was then dried at 37 °C for 1 h. Finally, the nitrocellulose membrane, conjugate pad, sample pad and the absorbent pads were assembled and cut into 4-mm strips.

### Evaluating the GICA strips

A positive serum sample was serially diluted with running buffer at a ratio of 1:10–1:320. The limit of detection of the GICA strip was determined using 100 μL of the sample, and 0.9% NaCl was used as the blank control. The results could be obtained by naked eyes after 5-10 min. Each test was repeated three times. Additionally, 41 serially-diluted samples collected from healthy men were tested to evaluate the specificity of the GICA strips.

Another 814 serum samples from patients suspected of harboring COVID-19 infections were collected from Huoshenshan Hospital, General Hospital of Central Threater Command of the PLA, the Sixth People’s Hospital of Shenyang, Peking Union Medical College Hospital, and Shijiazhuang Fifth Hospital in China. All samples were tested on the GICA strips, along with PCR tests on nasal/pharyngeal swab for comparison. PCRs were performed with officially approved real-time PCR (RT-PCR) kits.

## Results

### Acquisition of the six recombinant proteins

Through gene subcloning by overlapping extension PCR and ligation to the HEK 293 vector, the S1, S-RBD-mFc (containing the mouse Fc fragment) and S-RBD recombinant plasmids were constructed and verified by PCR and sequencing (Figure 1A). Following lipofection, transient expression in eukaryotic HEK293 cells and protein purification, highly pure rS1, rS-RBD-mFc and rS-RBD were obtained (Figure 1D).

Through gene subcloning and ligation to the pET vector, recombinant N, N1 and N2 plasmids were constructed and verified by PCR and sequencing (Figure 1B and C). The rN, rN1 and rN2 expression products from IPTG induction at 37°C in *E. coli* were identified in the culture supernatants and in the precipitates (Figure 1E and F), so the induction temperature was lowered to 20°C, which increased the protein in the supernatants significantly. The supernatant proteins were purified and used for subsequent studies (Figure 1E and F). That the HCoV-19 N proteins were present in the precipitates implies that their expression in *E. coli* may influence their accurate folding or conformation, because the viral N protein may be conformationally modified after transcription in human cells. Overall, six high-yield recombinant proteins from HCoV-19 with high purities were obtained, as summarized in Table 2.

**Figure 1.**
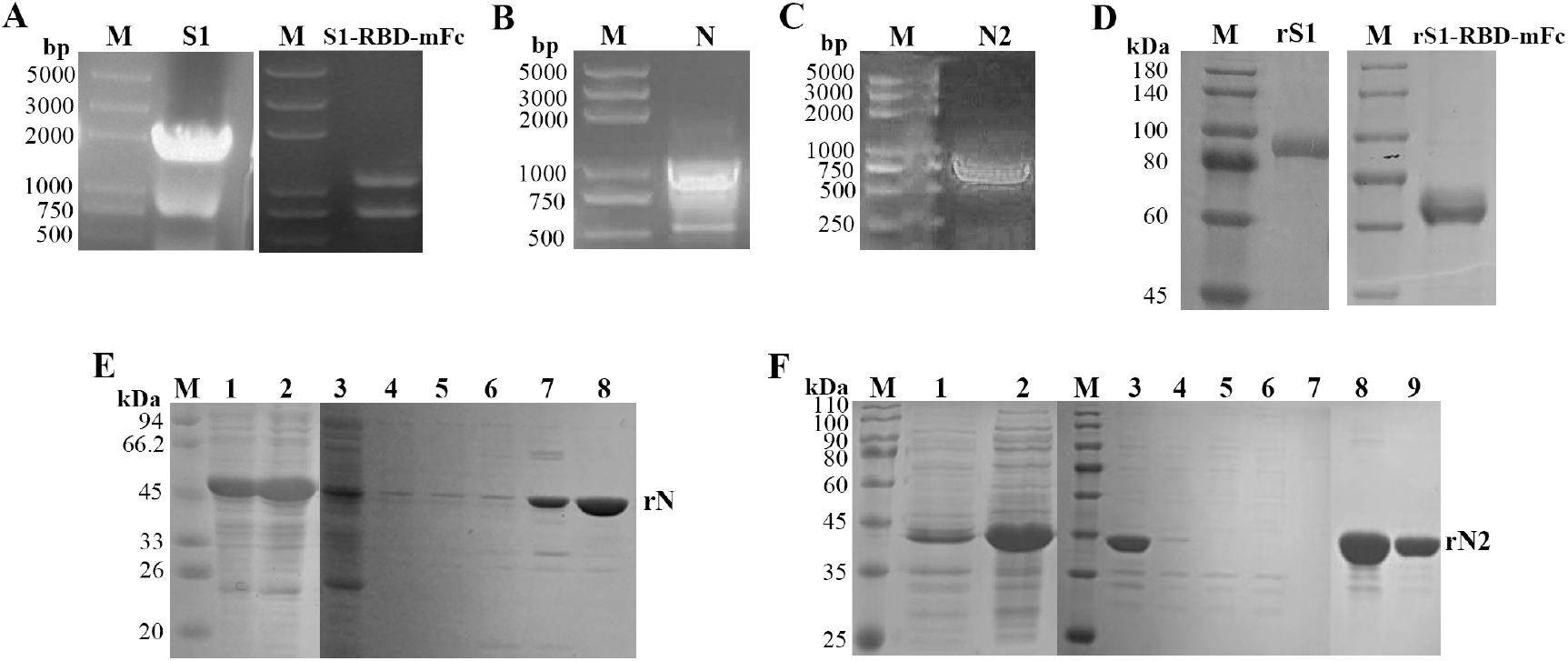
Gene fragments and purified recombinant HCoV-19 proteins analyzed by agarose electrophoresis and SDS-PAGE. (A) PCR identification of S1 and S-RBD-mFc gene fragments. (B) N gene fragment. (C) N2 gene fragment. (D) SDS-PAGE for purified rS1 and rS-RBD-mFc. (E) SDS-PAGE for rN. Lanes 1 and 2 are the precipitatant and supernatant, respectively, from crude *E. coli* extracts harboring the recombinant plasmid. Lanes 3–8: Ni Sepharose affinity column purification. Lane 3 shows unbound proteins. Lanes 4–7: proteins washed from the column using 10, 20, 60 and 80 mM imidazole. Lane 8 is rN eluted with 250 mM imidazole. (F) SDS-PAGE for rN2. Lanes 1 and 2 are the supernatant and precipitatant, respectively, from crude *E. coli* extracts harboring the recombinant plasmid. Lanes 3–9: Ni Sepharose affinity column purification. Lane 3 shows the unpurified supernatant proteins. Lanes 4–7: Proteins washed from the column using 10, 20, 20 and 50 mM imidazole. Lanes 8–9: rN eluted in 250 mM imidazole.

**Table 2.** Information of the prepared recombinant proteins.

| Protein | Tag<sup>a</sup> | Amino acid | MW<sup>b</sup> (kDa) | Expression | Strain<sup>c</sup> | Restriction site<sup>d</sup> |
| --- | --- | --- | --- | --- | --- | --- |
| **rS1-His** | His | 667 | 76.5 | Eukaryotic | HEK 293 | **EcoR I/**<br>**BamH I** |
| **rS-RBD-mFc** | His | 501 | 63 | Eukaryotic | HEK 293 | Afl II/Nhe I |
| **rS-RBD** | His | 259 | 30 | Eukaryotic | HEK 293 | Afl II/**BamH I** |
| **rN** | His | 428 | 45 | Prokaryotic<br>(pET28a) | *E.coli*<br>Rosetta | NcoI/XhoI |
| **rN1** | Sumo,<br>His | 273 | 47 | Prokaryotic<br>(pET28a) | *E.coli*<br>BL21 | **BamHI/**<br>**HindIII** |
| **rN2** | TrxA,<br>His | 210 | 43 | Prokaryotic<br>(pET32a) | *E.coli*<br>BL21 | NcoI/XhoI |

<sup>a</sup>Protein tags. Sumo and TrxA tags are 18 and 20 kDa, respectively.
<sup>b</sup> Molecular weight of the recombinant protein including the tags.
<sup>c</sup> Strains used for engineering and protein expression
<sup>d</sup> Restriction enzyme recognition sites

### Preliminary ELISA evaluation of recombinant proteins

Indirect ELISAs were used to preliminarily evaluate the six recombinant proteins. Using serum samples from seven healthy people as the negative controls, serial samples from two patients in the acute phase (<7 days after symptom onset) of COVID-19 were detected. For the S proteins, the coefficient of variation (CV) for the seven negative samples was 20%–30%. For the N proteins, the CV was 74%–92%, resulting in a high cutoff value.

For IgM detection, all the S proteins had lower backgroud values and higher OD values than those of the N proteins, as well as higher sentivities (1:800) (Figure 2A and 2C). For IgG detection, rS-RBD-mFc had the highest sensitivity (1:80), while the second highest sensivity (1:40) was attained by rS1, rS-RBD and rN (Figure 2B and 2D). Although with the same sensitivity and similar background, rS1 produced a higher OD value than rS-RBD. The better performance of rS-RBD-mFc when compared with rS-RBD may be related to the protein Fc fragments, which can increase the half-life and stability for the RBD protein fragment from the S protein. Overall, rS-RBD-mFc and rS1 were best suited for IgM and IgG detection.

The IgM-specific sensitivities for ELISA (from 1:100 to 1:800) are all higher than those for IgG (1:20–1:80), mostly because the positive serial samples came from patients during the acute phase of COVID-19 infection. Additionally, the antigenicity of full-length rN was higher than that of fragments rN1 and rN2, which is consistent with the findings from our previous study on SARS-CoV [8]. However, the higher sensitivity of rN1 over rN2 suggests that the antigenicity of the protein fragment at the NH_2_ terminus of the N protein was higher than that at the COOH terminus, which is the opposite result of our previous study on SARS-CoV [8].

**Figure 2.**
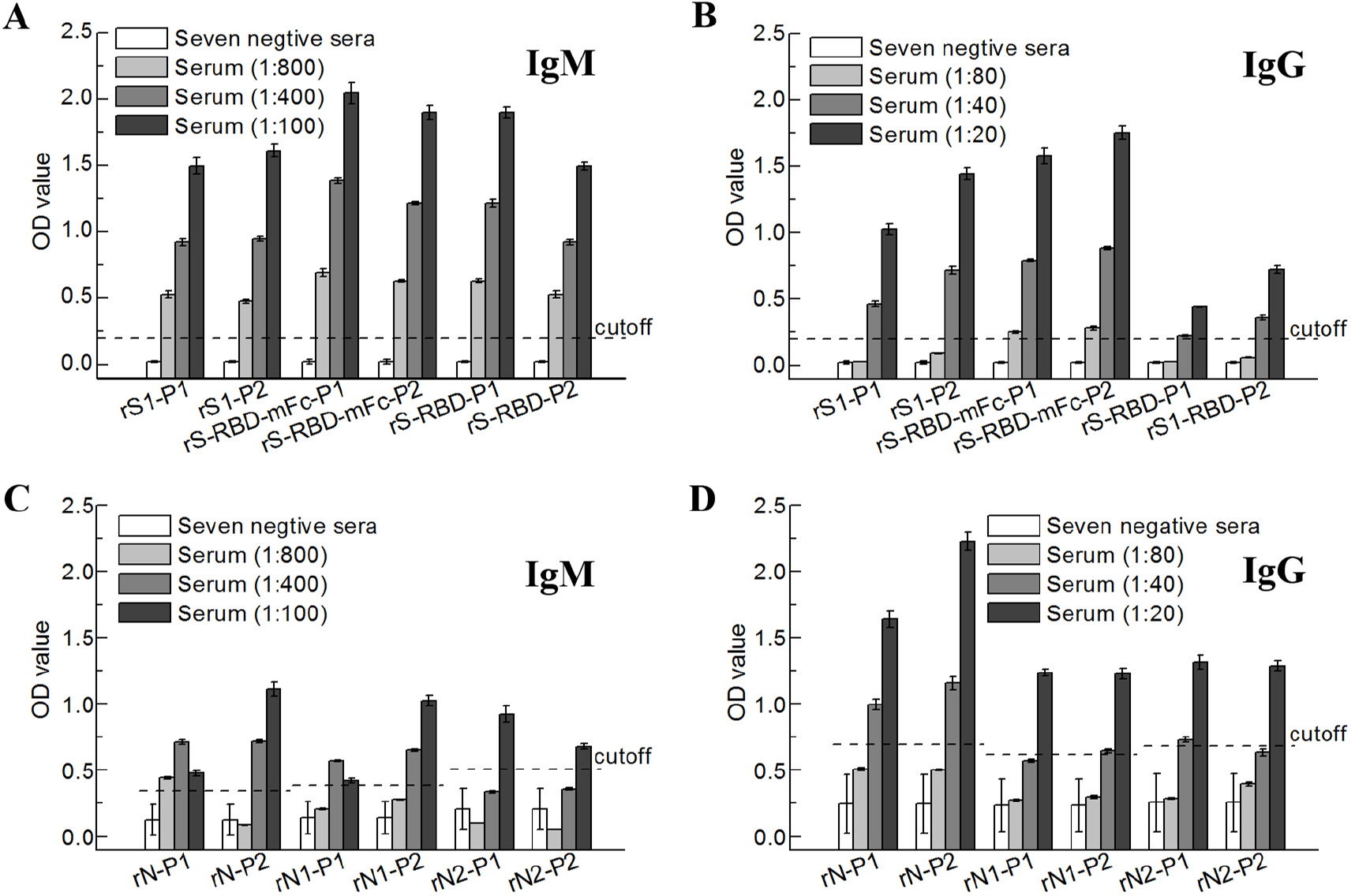
Evaluation of recombinant HCOV-19 proteins by indirect ELISA on serum samples. Serum samples from seven healthty people and two COVID-19 patients (P1 and P2) were used as negative controls and positive samples, respectively. (A) IgM detection in serum samples by the recombinant S1 domain protein fragment from the HCoV-19 spike protein (rS1), the receptor binding domain of the S protein (rS-RBD), and S-RBD ligated to the Fc fragment from mouse (rS-RBD-mFc) following their expression in a eukaryotic system. (B) IgG detection in serum samples using rS1, rS-RBD and rS-RBD-mFc proteins. (C) IgM detection in serum samples using full-length recombinant nucleocapsid protein (rN), an NH2 recombinant fragment from the N (rN1) or COOH (rN2) termini following their expression in a prokaryotic system. (D) IgG detection in serum samples using rN, rN1 and rN2 proteins. Dashed lines, cutoff values.

### Recombinant protein evaluation by GICA tests

As shown in Figure 3, a double-antigen sandwich-format GICA strip developed with recombinant rS-RBD and rS1 proteins was used to detect total antibodies (IgM and IgG) against HCoV-19 in serum samples. Positive results were observed at a 1:160 dilution of the patient serum, indicating that the GICA strip was able to detect low-titer antibodies in serum. Moreover, several samples from patients infected with influenza A, influenza B, respiratory syncytial virus, *Mycoplasma pneumoniae*, and *Chlamydia pneumoniae* were all tested by GICA. No cross-reactions were observed with these samples. An additional 41 serum samples from healthy people were all negatively detected by the GICA strips, demonstrating good specificity for the rS-RBD- and rS1-based serological methods.

**Figure 3.**
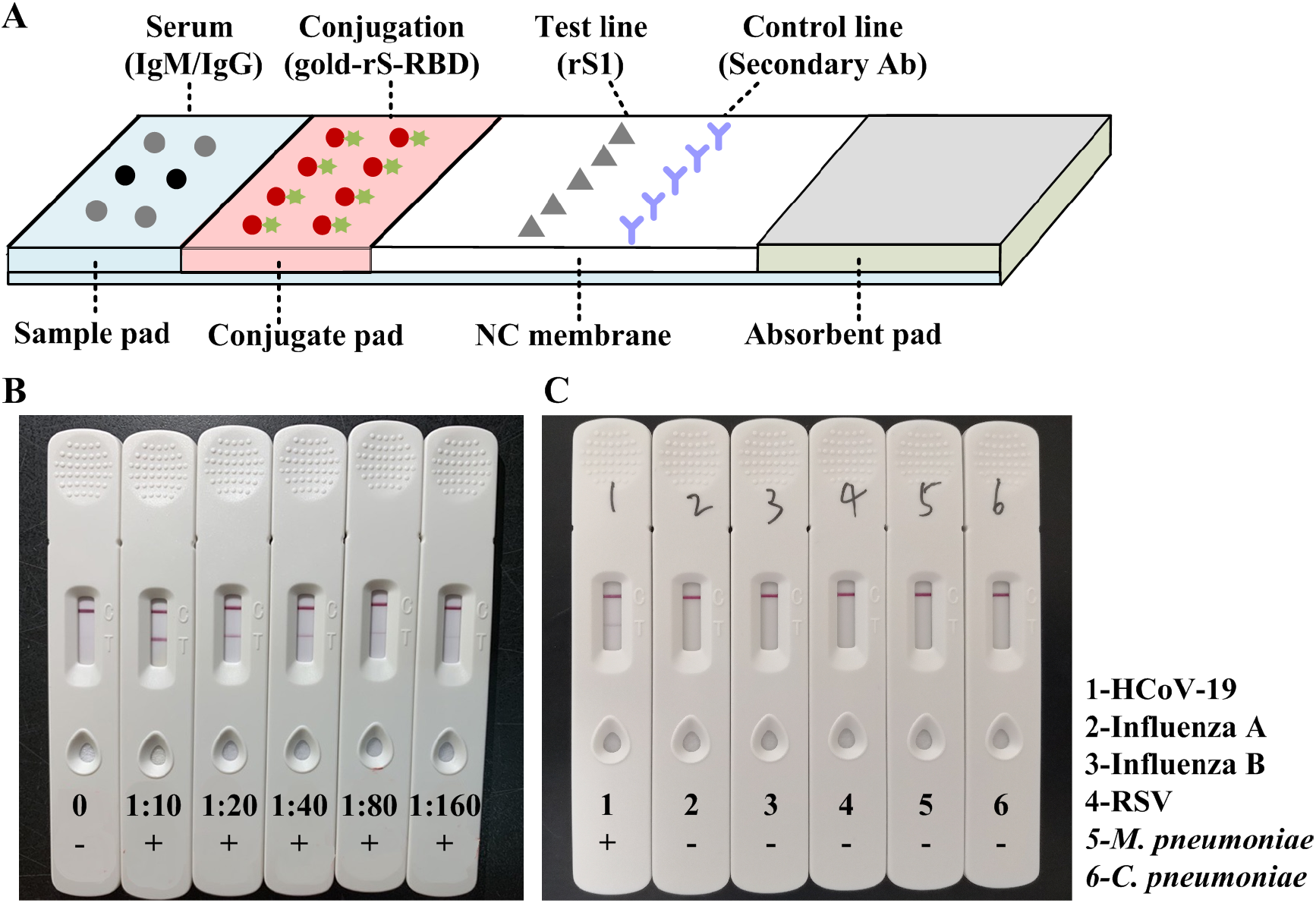
(A) Schematic illustration of the sandwich-format GICA strip for IgM and IgG antibody detection against HCoV-19. (B) Lower detection limit of the GICA strip, as determined with serial dilutions of the positive serum. (C) Cross-reaction testing of the GICA strip with other respiratory diseases.

### GICA strip performances on serum samples

Altogether, 814 serum samples from HCoV-19-suspected patients were tested on GICA strips, including 122 patients confirmed to be RT-PCR positive, 660 confirmed to be RT-PCR negative, and 32 confirmed to be RT-PCR negative but clinically diagnosed by computed tomography (CT). The positive coincidence and negative coincidence rates between GICA and RT-PCR were 86.89% and 99.39%, respectively (Table 3). Moreover, among the 32 RT-PCR negative clinically diagnosed samples, 21 (65.63%) were detectable by GICA. This shows that GICA has a high coincidence rate with RT-PCR tests and also complements the PCR negative, clinically diagnosed cases, providing a potentially powerful serological tool for diagnosing COVID-19 in patients.

**Table 3.** GICA strip results of the serum samples.

| Group | Total | GICA detection results (%) |  |
| --- | --- | --- | --- |
|  |  | Positive | Negative |
| **RT-PCR Positive** | 122 | 106 (86.89%) | 16 (13.11%) |
| **RT-PCR Negative** | 660 | 4 (0.61%) | 656 (99.39%) |
| **Clinically confirmed** | 32 | 21 (65.63%) | 11 (35.37%) |

## Discussion

Rapid detection methods are urgently required for the early diagnosis of patients with fever [10, 12, 13], and for pre-symptomatic or asymptomatic carriers infected with HCoV-19 [12, 14]. However, nucleic acid-based molecular diagnosis tools (e.g., real-time PCR and loop-mediated isothermal amplification), depend on there being a sufficient viral load in the patient’s upper respiratory tract and reasonable sample quality. In the early infection stages, human antibodies, especially IgM, can reach a high level during the immune response to viral invasion. Therefore, serological testing is important for early COVID-19 diagnosis, especially for patients with undetectable viral RNA [10].

Selecting suitable recombinant HCoV-19 proteins for reliable serological assays is crucial. Some cross-reactive N proteins from another coronavirus were serologically investigated to meet the rapidly developing HCoV-19 emergency [9, 15]; however, the divergent receptor binding protein spike gene in other CoVs [9], as well as the high antigenicity of the receptor binding domain of HCoV-19 [3] emphasized the necessity to research HCoV-19 antigens more comprehensively. Consequently, in the present study, we prepared six recombinant proteins, including three S proteins (rS1, rS-RBD, rS-RBD-mFc) and three N proteins (rN, rN1, rN2). Because the S protein is a transmembrane protein, three S protein fragments were prepared using a eukaryotic expression system, while three N proteins were prepared using a prokaryotic expression system. Preliminary evaluation by indirect ELISA revealed that the three S proteins were superior in performance to the three N proteins. Thus, it can be inferred that the eukaryotically expressed HCOV-19 recombinant S protein is suitable for detection of antibodies from COVID-19 patients for early diagnosis, and is superior to the N protein prepared by prokaryotic expression. However, the antigenicity of S proteins and that of the N proteins could not be confirmed in this study, because the influence of prokaryotic expression method used to produce the N proteins is difficult to predict. The prokaryotically expressed recombinant N proteins were all insoluble precipitates, especially rN2, making it likely that their natural conformations were lost or they were not normally modified. Thus, the N protein’s antigenicity should be studied after expression in a eukaryotic system in future.

GICA, a simple and rapid serological method, is especially suitable for timely diagnosis and large-scale sample screening. A recent study showed that the sensitivity of the total antibody (IgM and IgG) test is higher than that of the IgM or IgG test [16]. Therefore, we developed a sandwich-format GICA with rS1 and rS-RBD-mFc to detect total antibodies in patient serum. The GICA tests were highly sensitive (86.89%) and specific (99.39%), which should help with diagnosing COVID-19 in RT-PCR-negative patients who are clinically confirmed COVID-19 positive by CT. The detection sensitivity will be improved if we eliminate the sera from patients within the 5 days of symptoms onset.

## Conclusions

In the study, six recombinant HCoV-19 proteins were prepared and selected. It showed that the eukaryotically-expressed spike proteins (rS1and rS-RBD-mFc) are more suitable than the prokaryotically expressed nucleocapsid proteins for HCoV-19 serological diagnosis. The provided recombinant proteins could be used for development of serological diagnostic tools for COVID-19 infections. Here, A sandwich-format GICA strip with the recombinant proteins was developed to detect total antibodies (IgM and IgG) against HCoV-19. The assay was shown to be a rapid (10 min), simple, and highly sensitive diagnostic tool, providing a potentially powerful serological method for diagnosing COVID-19.

## Data Availability

The raw/processed data required to reproduce these findings cannot be shared at this time as the data also forms part of an ongoing study.

## Notes

## Acknowledgments

We thank the clinical study investigators and volunteers. We thank for our collaborators in the Laboratory of Protein Project, Beijing Institute of Biotechnology, for providing the eukaryotic vectors.

## Potential conflicts of interest

All authors: No reported conflicts of interest. All authors have submitted the ICMJE Form for Disclosure of Potential Conflicts of Interest.

**No part of the information has previously been presented in any meetings**.

## Correspondence

Y. Z, Dr., Beijing Institute of Microbiology and Epidemiology, No. 20, Dongdajie street, Fengtai district, Beijing 100071, P. R. China

C. L., Mr., Beijing Key Laboratory of POCT for Bioemergency and Clinic (No. BZ0329), Beijing 100071, P. R. China

S.L., Dr., Wuhan University School of Health Sciences, Wuhan 430071, P. R. China

